# Awareness and knowledge of interventional radiology among medical students at an Indian institution

**DOI:** 10.1101/19010066

**Authors:** Deepsha Agrawal, Michael Alan Renfrew, Sulove Singhal, Yash Bhansali

**Affiliations:** County Durham and Darlington NHS Foundation Trust, UK; University of Edinburgh, UK; Pt JNM Medical College, India; Syracuse University, NY, USA

## Abstract

**Purpose:** Interventional radiology (IR) is a novel and evolving sub-specialty that encompasses image guided diagnostic and therapeutic procedures. With the advent of new imaging techniques and an increasing demand of minimally invasive procedures IR continues to grow as a core component in medical and surgical therapeutics. Radiology teaching is a part of medical undergraduate curriculum, however, the medical undergraduate cohort lacks exposure to IR principles, methods and techniques.

The purpose of this study is to determine the knowledge and awareness of IR among medical students in a single university in India.

**Materials and Methods:** Anonymous electronic surveys were sent to 350 medical students of Pt. JNM Medical College, Raipur, India. Each survey comprised of questions assessing knowledge and exposure to IR. A total of 70 students (20%) responded.

**Results:** 85.7% of respondents believed that radiologists have a role in diagnostic as well as therapeutic interventions, however, 60% of students cited a very poor/poor knowledge of IR. A larger part, 91.5%, stated that they would be interested in IR based teaching delivered as a part of their undergraduate teaching program. Those who knew at least one interventional radiology technique were 1.51 (95% CI: 1.02 - 2.22; p < 0.05) times more likely to consider it as a career.

**Conclusion:** Medical Students demonstrate a poor knowledge of IR. This corresponds to a limited and inconsistent exposure to IR in medical schools. The study suggests that there is a need to deliver an IR based curriculum in medical undergraduate teaching in India.

## Introduction

Interventional radiology (IR) has experienced an unprecedented growth in recent years. Having evolved from limb saving angioplasties in 1964 to over 50 therapeutic procedures today, the subspecialty of IR continues to flourish. A look at the Google ngram for phrase ‘interventional radiology’ aptly and interestingly represents the flourishing practice of IR. The graph begins at a frequency of 0.0000000092%, being at the baseline of 0.00000000% until then, and exponentially rises up to a frequency of 0.0000057220% in 2008 [Figure 1]. This graph charts the frequency of ‘interventional radiology’ search string in Google’s text corpora in English.

**Figure 1.**
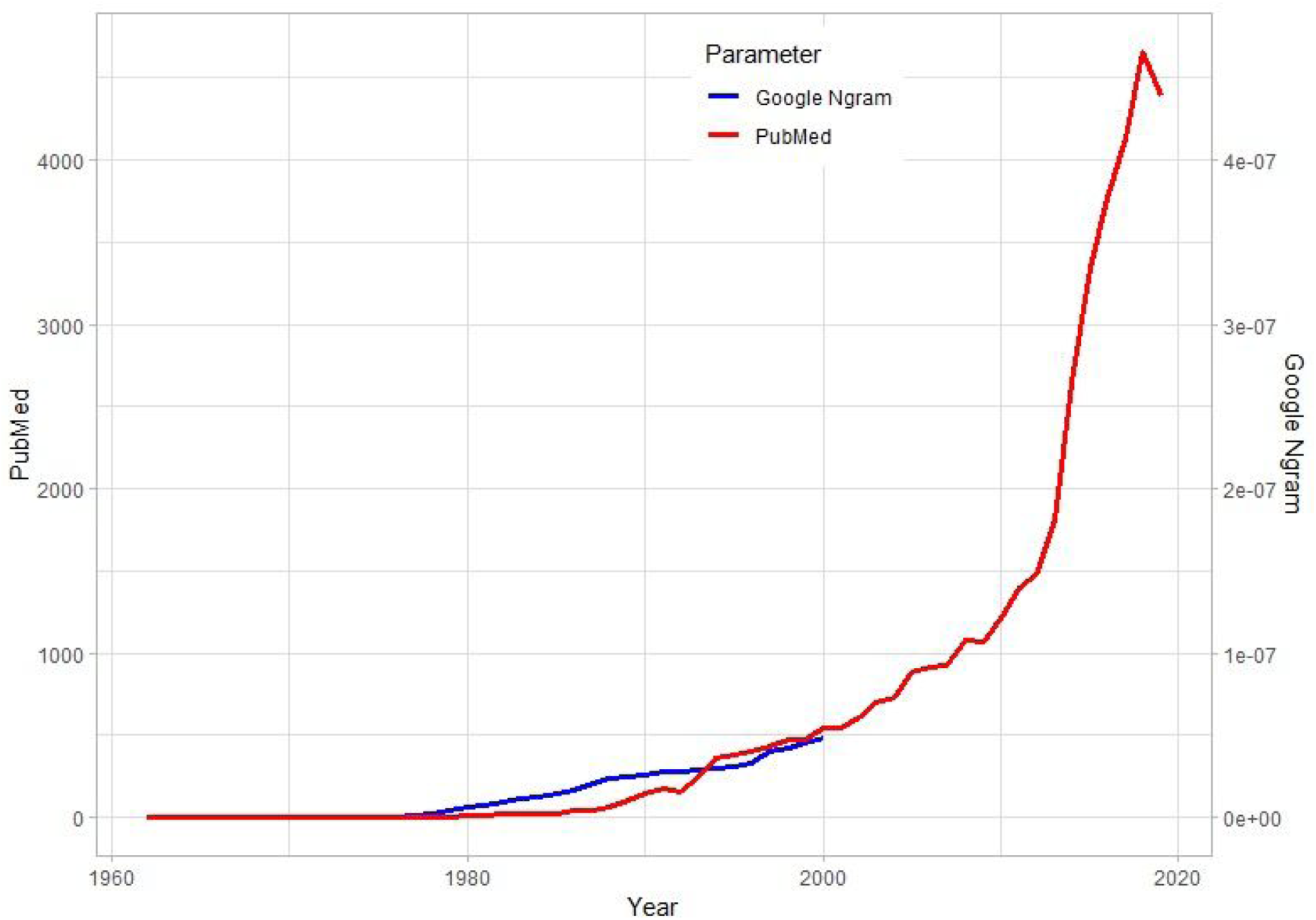
A graphical representation of texts indexed under word string ‘interventional radiology’ on Google English corpora and PubMed between 1960 and 2019.

A key World Health Organization (WHO) report entitled ‘Efficacy and radiation safety in interventional radiology’, published in 2000, also concluded that IR has a growing scope of practice in treating diseases of cardiovascular and non-vascular origin in both developed and developing countries [1]. However, due to only recent accreditation of IR as a sub-specialty, modules for IR based teaching have not been amalgamated into undergraduate medical curriculum. In several studies across different countries, medical students have demonstrated a poor understanding of IR, owing to suboptimal teaching and exposure levels [2-10].

As technical means to provide minimally invasive interventions with better outcomes continue to grow, it is of paramount importance to train the future workforce. The Cardiovascular and Interventional Society of Europe (CIRSE) [11], British Society of Interventional Radiology (BSIR) [12] and the Society of Interventional Radiology (SIR) [13], and have been continuously working towards addressing a lack of IR knowledge and awareness among medical students.

There is a lack of formal evidence from India, however, there have been reports of a steady growth in technology and therapeutic radiology procedures. This study is derived from a survey presented to second, pre-final and final year medical students in an Indian university. It was conducted to appraise the current understanding and perception of IR among medical students. The results depict medical students’ awareness under the present curriculum and extend into their perception of radiation exposure in IR.

## Material and Methods

An electronic survey with 12 questions was designed based on the studies conducted by Gregorio et al. [4] and Leong et al. [6]. This electronic survey was distributed among 350 medical students of Pt. Jawarharlal Nehru Memorial Medical College, Raipur, India. The survey was entirely electronic, anonymous and participation was voluntary. Responses were collected during July 2019, starting from 1 July 2019 to 31 July 2019.

The questions demanded a mandatory response on year of medical school. All students from the first year of medical school and internship were excluded from the study. Questions were based on a single choice between ‘Yes’ and ‘No’. Some questions took into account the level of knowledge/awareness and were marked on a range of very poor to very good or strongly disagree to strongly agree. Questions relating to understanding of procedures accepted multiple responses against designated procedures on the survey form. The form allowed students to enter an expansive answer regarding their concerns about radiation exposure in the practice of IR.

## Statistical Analysis

Responses collected from the survey were collated in a spreadsheet program (Excel, Microsoft 2010) and analyzed using statistical software (R-3.6.1 for windows 10). Data were tested using Fisher’s exact test. P value was established at 0.038 (P<0.05).

## Results

A total of 70 (20%) out of potential 350 students responded to the survey. These participants were distributed across pre-clinical and clinical year-16/22.9% in pre-clinical (2nd year) and 54/77.1 in clinical (pre-final and final year). Most of the students-85.7% knew that radiology could be diagnostic as well as therapeutic. However, the number slightly dipped down to 78.6% when asked whether they understand what IR is [Figure 2]. Of those who stated they discerned what is IR, a 49% of majority indicated that it was through classroom teaching. The remainder answered as follows: 20% at the hand of teachers/senior colleagues outside of classroom teaching; 12% via internet resources; 9% through social media; 5% by means of clinical setting and interestingly 5% through the medium of medical TV shows [Figure 3].

**Figure 2.**
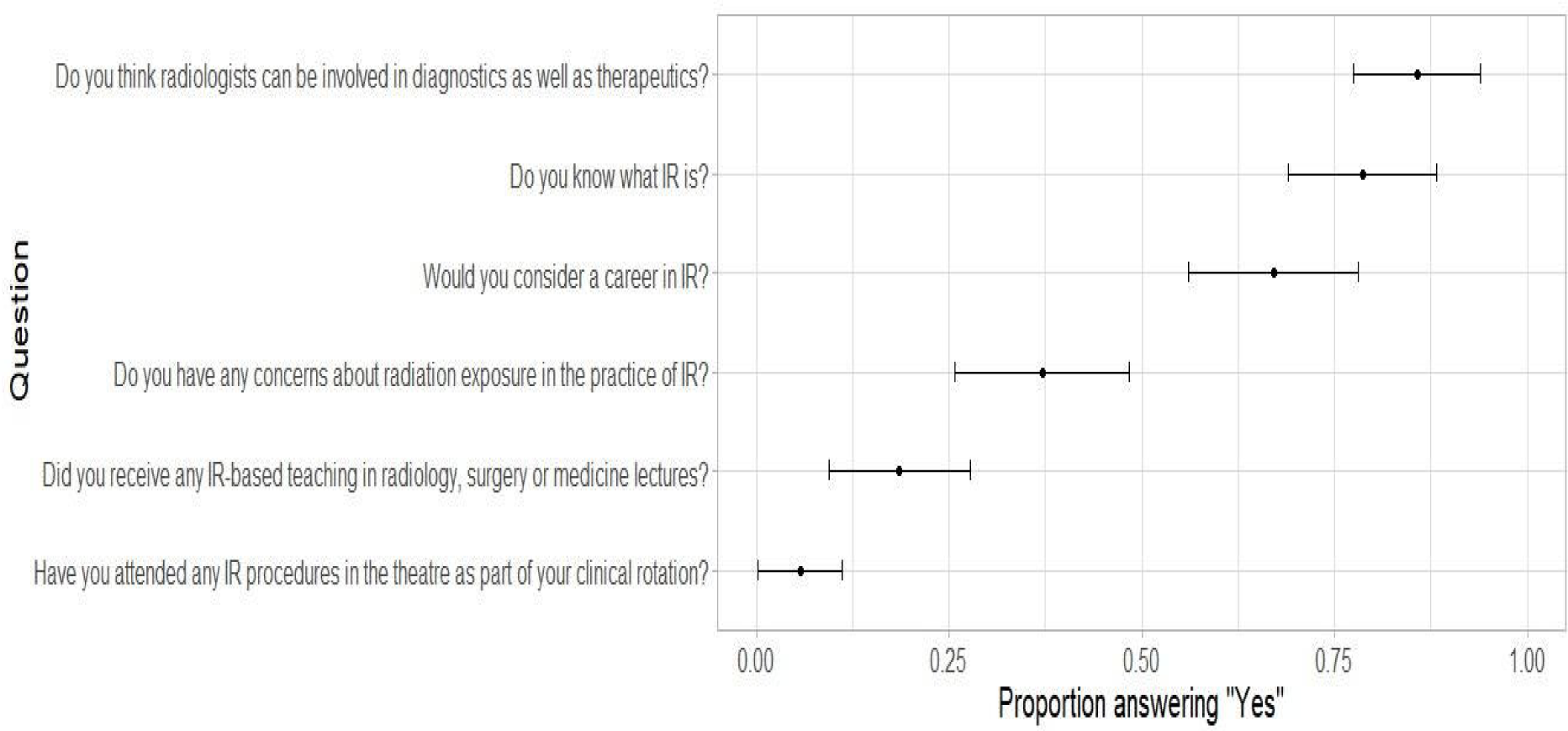
N=70. An analysis of survey questions (y-axis) and responses (x-axis) depicted on a 95% CI.

**Figure 3.**
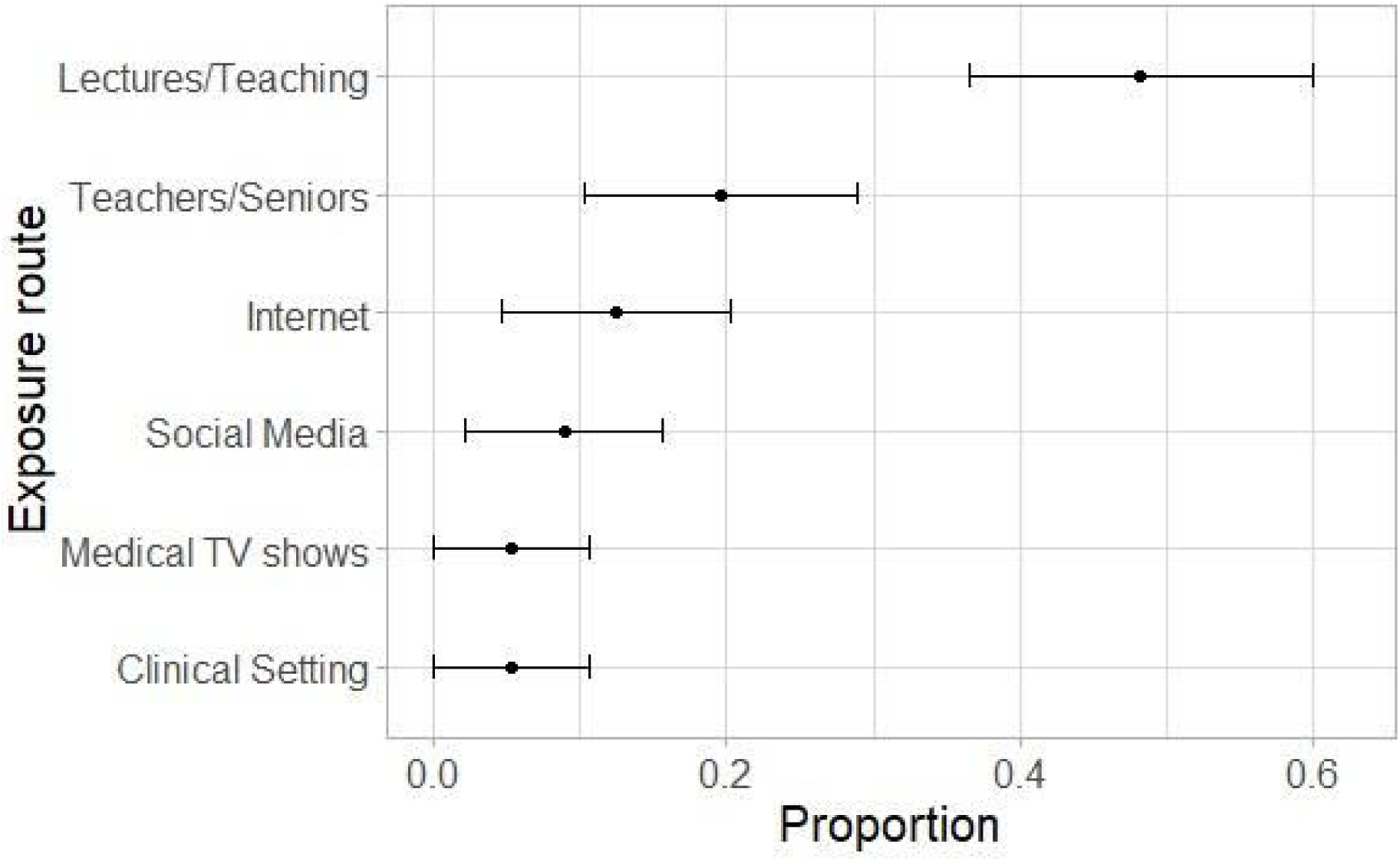
N= 59. Most students attribute awareness of IR through lectures/ teaching in medical schools.

Upon being asked to self-evaluate the knowledge of IR as compared to other medical specialties, most students admitted poor to very poor knowledge. 41.4 and 18.6% students indicated poor and very poor knowledge, respectively against a 27.1 % reporting a fair level of knowledge of IR. Only 7.1 and 5.7% students agreed to having good and very good knowledge, respectively [Figure 4].

**Figure 4.**
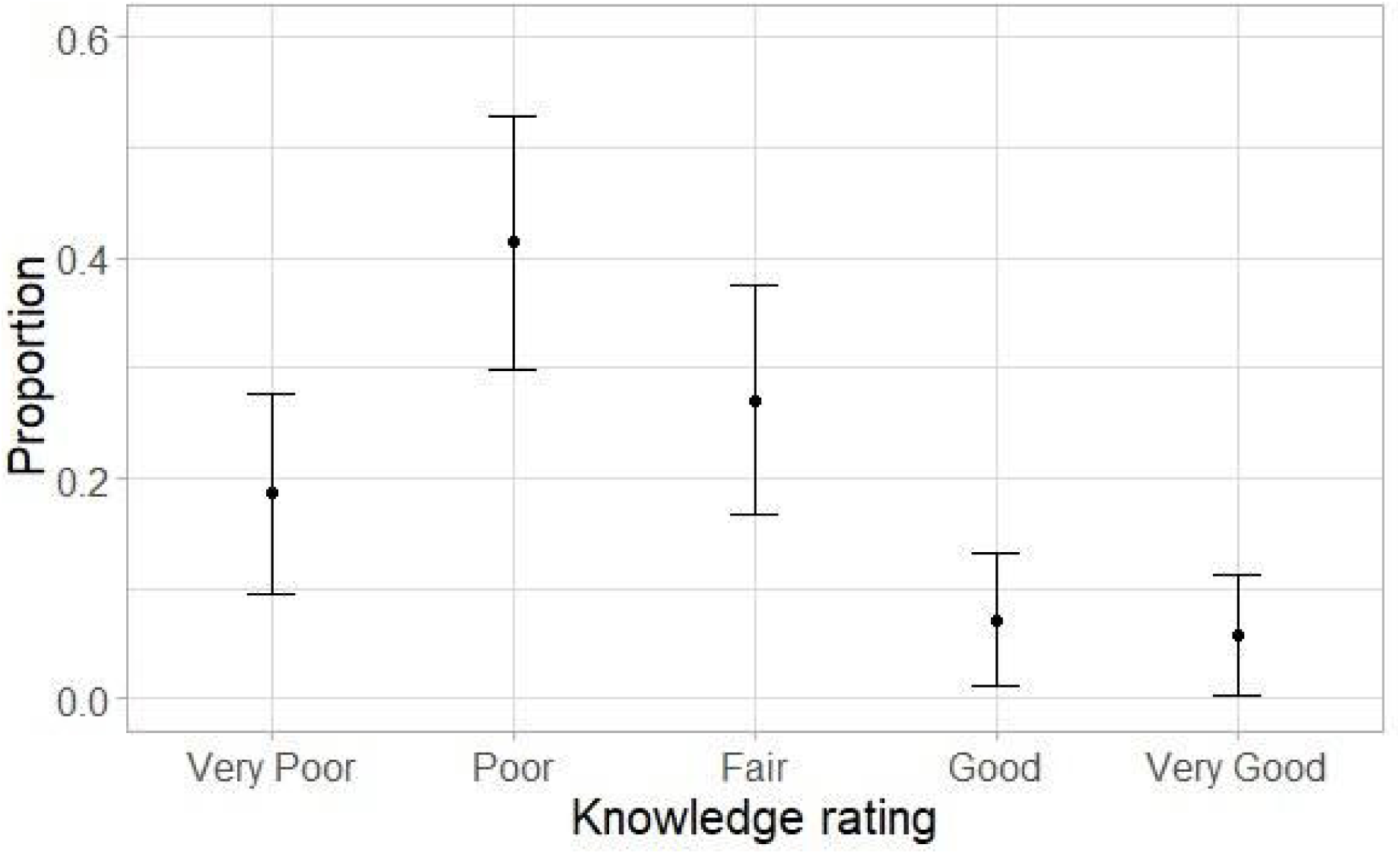
N=70. A proportion of 0.41 and 0.18 estimate their IR knowledge as ‘poor’ and ‘very poor’ respectively.

The next question surveyed if the students had received any introductory teaching on IR in medicine or surgery classes. A major fraction of students, 81.4%/57, said no while a smaller fraction of 18.6%/13answered yes. Out of the 13 students who answered yes, 10 expressed where they received this introductory teaching. Most (70%) of this was indicated as teaching in dedicated radiology classes. However, on being surveyed on the scope of IR procedures the respondents attended as part of their clinical rotation, 94.3% registered a ‘No’ as an answer.

Regarding the awareness of the kind of procedures interventional radiologists do in daily practice, the answers ranged from don’t know to endovascular coil embolization. 41.4% said they had no idea, 5.7 answered incorrectly and 52.9% answered correctly choosing among endovascular coil embolization, ultrasound guided biopsies, balloon angioplasty, uterine fibroid embolization and percutaneous nephrostomy.

As a concluding assessment to overall perception of medical students, it was asked if they feel there should be more IR-orientated teaching as a part of the undergraduate curriculum. Most of the students, 91.5%, marked agree (72.9%) and strongly agree (18.6%) [Figure 5].

62.9% of respondents also acknowledged a concern about radiation exposure in IR, nonetheless 67.1.% said they would consider a career in IR.

**Figure 5.**
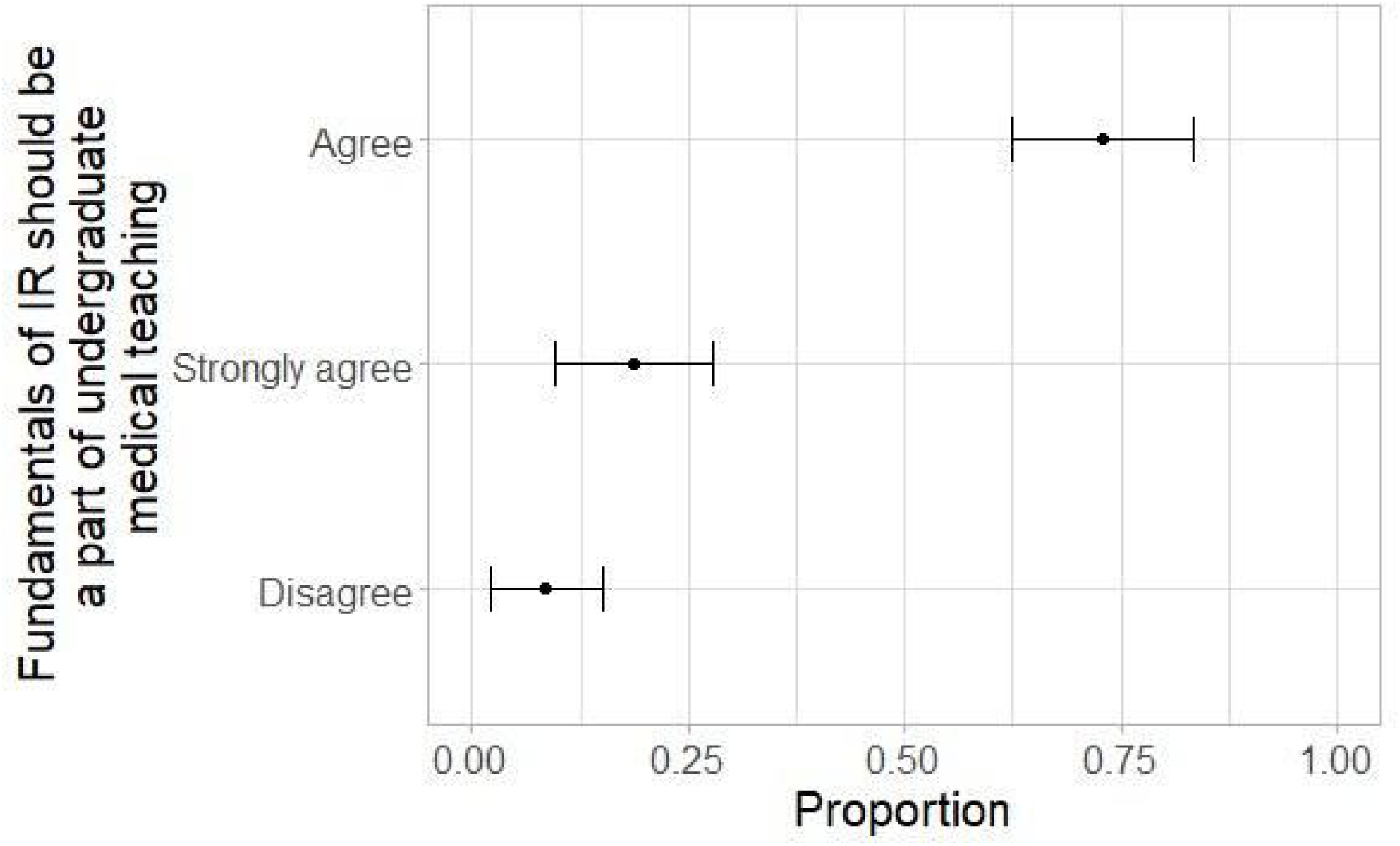
N=70. Majority of medical students agree to integrating IR in undergraduate medical teaching.

## Discussion and Conclusion

The most important finding of the study was that those who knew at least one interventional radiology technique were 1.51 (95% CI: 1.02 - 2.22; p < 0.05) times more likely to be considering it as a career. This suggests that teaching more techniques may engender interest in the field, although the effects may be simply correlated: those who are already interested may make more of an effort to learn techniques.

The other important finding is that the students expressed a lack of knowledge and insufficient exposure to IR in their undergraduate years.

Interventional radiology is one of the most dynamic fields in medicine and as it continues to evolve, there is a need for an undergraduate curriculum appropriate for medical schools. An under-representation of IR in undergraduate modules could have a direct impact on both, the choice of IR as a career and for the future practice of medicine irrespective of their specialty This, in turn, suggests a need to revise the current radiology curriculum and clinical rotations to introduce IR at an early stage of medical education.

It is recommended that medical students receive a curriculum based teaching in view of the common medical conditions and their image-guided interventional treatments [Table 1]. Enriching the undergraduate cohort with the fundamental principles underpinning IR may further a critical understanding of it [14,15,16,17]. This curriculum would ideally be amalgamated into medical school and internship assessments.

**Table 1:** Principle IR subjects, presently.

| Condition | Treatment |
| --- | --- |
| Peripheral Arterial Disease | Angioplasty and stenting |
| Aneurysm | Endovascular Repair (EVAR) |
| Venous Thromboembolic Disease | Catheter-directed thrombolysis, balloon angioplasty, or stenting. Alternatively, IVC filter |
| Uterine Fibroids | Uterine artery embolization |
| Benign prostate hyperplasia (BPH) | Prostate artery embolization |
| Biopsies and drainage | Image-guided access using US, CT or fluoroscopy |
| Vascular Malformations | Sclerotherapy |
| Interventional Oncology | Radiofrequency ablation, cryoablation, chemoembolization |
| Stroke | Intra-arterial thrombolysis treatment |
| Portal Hypertension | Transjugular intrahepatic portosystemic shunt (TIPS) procedure |

The cardiovascular and interventional society of Europe (CIRSE) recommends that the medical students IR curriculum should deliver the following [11]:

- The basis of interventional radiology and its historical context
- Image guidance for interventional procedures
- Knowledge of radiation protection guidelines for interventional procedures
- Knowledge of the legislation relating to the use of interventional radiology in clinical practice

The present study has several limitations. Firstly, a relatively small number of students were surveyed. Secondly, these students belonged to the same university, therefore had similar academic environment. Moreover, a lot of respondents indicated IR teaching outside of medical school, which shifts the knowledge positively, however, is not delivered as a part of undergraduate medical curriculum. Therefore, further studies are needed to validate our proposition.

In conclusion, this study demonstrates that medical undergraduate students in India have a poor understanding of IR due to limited exposure to the sub-specialty. An intervention, in the form of introducing IR in medical undergraduate teaching may have a positive impact on the knowledge and skills of medical students and a critical understanding of the scope of practice in IR.

It is acknowledged that the inception of these developments will be a challenge; particularly facets such as working in collaboration with the regulatory bodies to introduce a curriculum and a delivery outlook.

## Data Availability

not applicable

## Notes

### Competing Interest Statement

The authors have declared no competing interest.

### Funding Statement

No funding received

### Author Declarations

All relevant ethical guidelines have been followed and any necessary IRB and/or ethics committee approvals have been obtained.

Any clinical trials involved have been registered with an ICMJE-approved registry such as ClinicalTrials.gov and the trial ID is included in the manuscript.

## References

1. Worldcat.org. (2019). Efficacy and radiation safety in interventional radiology. (Book, 2000) [WorldCat.org]. [online] Available at: https://www.worldcat.org/title/efficacy-and-radiation-safety-in-interventional-radiology/oclc/45947497.

2. O’Malley, L. and Athreya, S. (2012). Awareness and Level of Knowledge of Interventional Radiology among Medical Students at a Canadian Institution. Academic Radiology, 19(7), pp.894–901.

3. Atiiga, P., Drozd, M. and Veettil, R. (2017). Awareness, knowledge, and interest in interventional radiology among final year medical students in England. Clinical Radiology, 72(9), pp.795.e7–795.e12.

4. de Gregorio, M., Guirola, J., Sierre, S., Serrano-Casorran, C., Gimeno, M. and Urbano, J. (2018). Interventional Radiology and Spanish Medical Students: A Survey of Knowledge and Interests in Preclinical and Clinical Courses. CardioVascular and Interventional Radiology, 41(10), pp.1590–1598.

5. Commander, C., Pabon-Ramos, W., Isaacson, A., Yu, H., Burke, C. and Dixon, R. (2014). Assessing Medical Students’ Knowledge of IR at Two American Medical Schools. Journal of Vascular and Interventional Radiology, 25(11), pp.1801–1807.e5.

6. Leong, S., Keeling, A. and Lee, M. (2009). A Survey of Interventional Radiology Awareness Among Final-Year Medical Students in a European Country. CardioVascular and Interventional Radiology, 32(4), pp.623–629.

7. Ghatan, C., Kuo, W., Hofmann, L. and Kothary, N. (2010). Making the Case for Early Medical Student Education in Interventional Radiology: A Survey of 2nd-year Students in a Single U.S. Institution. Journal of Vascular and Interventional Radiology, 21(4), pp.549–553.

8. Nissim, L., Krupinski, E., Hunter, T. and Taljanovic, M. (2013). Exposure to, Understanding of, and Interest in Interventional Radiology in American Medical Students. Academic Radiology, 20(4), pp.493–499.

9. Foo, M., Maingard, J., Phan, K., Lim, R., Chandra, R., Lee, M., Asadi, H., Kok, H. and Brooks, M. (2018). Australian students’ perspective on interventional radiology education: A prospective cross-institutional study. Journal of Medical Imaging and Radiation Oncology, 62(6), pp.758–763.

10. Ojha, U., Mohammed, R. and Vivekanantham, S. (2017). Should there be greater exposure to interventional radiology in the undergraduate curriculum? Advances in Medical Education and Practice, Volume 8, pp.791–795.

11. Cirse.org. (2019). [online] Available at: https://www.cirse.org/wp-content/uploads/2018/07/CIRSE_IR_Curriculum_for_Medical_Students.pdf [Accessed 9 Oct. 2019].

12. Bsir.org. (2019). [online] Available at: https://www.bsir.org/media/resources/UK_Undergraduate_Curriculum_for_IR_2014.pdf.

13. Sirweb.org. (2019). Society of Interventional Radiology-Society of Interventional Radiology. [online] Available at: https://www.sirweb.org/.

14. Shaikh, M., Shaygi, B., Asadi, H., Thanaratnam, P., Pennycooke, K., Mirza, M. and Lee, M. (2015). The Introduction of an Undergraduate Interventional Radiology (IR) Curriculum: Impact on Medical Student Knowledge and Interest in IR. CardioVascular and Interventional Radiology, 39(4), pp.514–521.

15. Alexander, E., Machan, J. and Ahn, S. (2015). Early Introduction of IR to Premedical and Medical Students: Initiatives at a Single U.S. Institution. Journal of Vascular and Interventional Radiology, 26(3), pp.439–442.

16. Lee, A. and Lee, M. (2017). Teaching IR to Medical Students: A Call to Action. CardioVascular and Interventional Radiology, 41(2), pp.203–205.

17. Goldman, D., Magnowski, A., Rochon, P., Bream, P., Kondo, K., Peters, G., Martin, J. and Fischman, A. (2018). The State of Medical Student Teaching of Interventional Radiology: Implications for the Future. Journal of the American College of Radiology, 15(12), pp.1761–1764.

